# Effect of an intensive lifestyle intervention on the structural and functional substrate for atrial fibrillation in people with metabolic syndrome

**DOI:** 10.1101/2023.08.02.23293550

**Authors:** Xavier Rossello, Raúl Ramallal, Dora Romaguera, Ángel M. Alonso-Gómez, Alvaro Alonso, Lucas Tojal-Sierra, Carlos Fernández-Palomeque, Miguel Ángel Martínez-González, María Garrido-Uriarte, Luis López, Agnes Díaz, Olatz Zaldua-Irastorza, Amit J. Shah, Jordi Salas-Salvadó, Montserrat Fitó, Estefania Toledo

## Abstract

**Aims:** To evaluate the effect of an intensive lifestyle intervention (ILI) on the structural and functional cardiac substrate of atrial fibrillation (AF) in overweight or obese people with metabolic syndrome (MetS).

**Methods:** Participants of the PREDIMED-PLUS trial (n=6874) were randomised 1:1 to an ILI program based on an energy-reduced Mediterranean diet, increased physical activity, and cognitive-behavioural weight management, or to a control intervention of low-intensity dietary advice. Left atrial (LA) strain, function, and volumes were evaluated by a core echocardiography lab in 534 participants at baseline, 3-year and 5-year follow-up. Mixed models were used to evaluate the effect of the ILI on LA structure and function.

**Results:** In the subsample, baseline mean age was 65 years (SD 5 years), and 40% of the participants were women. Over the 5-year period, both groups experienced worsening of LA structure and function, with increases in LA volumes and stiffness index and decreases in LA longitudinal strain, LA function index and LA emptying fraction over time. Changes in the ILI and control group were not significantly different for any of the primary outcomes (LA emptying fraction: -0.95% (95%CI -0.93, -0.98) in control group, -0.97% (95%CI -0.94, -1.00) in ILI group, p_between groups_=0.80; LA longitudinal strain: 0.82% (95%CI 0.79, 0.85) in control group, 0.85% (95%CI 0.82, 0.89) in ILI group, p_between groups_=0.24) or any of the secondary outcomes.

**Conclusions:** In overweight or obese people with MetS, an ILI had no impact on the underlying structural and functional left atrial substrate measurements associated with AF risk.

## INTRODUCTION

Atrial fibrillation (AF) is the most prevalent cardiac arrhythmia, and a risk factor for stroke, heart failure, dementia, and mortality. ^1,2^ AF is a progressive disease, with many patients advancing over time from subclinical states (changes in the atrial substrate) to clinical forms of the arrhythmia (paroxysmal, persistent and then permanent AF).^3^ Before AF occurs, several adaptive functional and structural changes may take place in the left atrium (LA).^3^ Echocardiography is the foremost imaging modality employed to study the structure and function of the left atrium. Atrial structural anatomy can be evaluated using either two-dimensional (2D) or three-dimensional transthoracic echocardiography (TTE). For early detection of functional changes before anatomical changes become evident, 2D speckle-tracking echocardiography is a more sensitive technique. Strain imaging provides reliable information on myocardial deformation by estimating differences in myocardial velocities. LA strain is not commonly obtained, yet growing literature has supported its role in numerous outcomes.^4^ These atrial changes, as well as the likelihood to progress to AF, are mostly determined by the presence of concomitant risk factors. Whether or not risk factor control may influence pathological LA changes predisposing to AF is not known, but important when considering public health prevention programming.

Both metabolic syndrome (MetS) and increased body weight are major risk factors for AF development and progression, since they have an impact on cardiovascular hemodynamics as well as on cardiac function and structure.^5–7^ Several studies have shown that lifestyle changes controlling such risk factors may play a role in AF prevention.^3,8^ In this line, a meta-analysis of randomised and observational studies reported that weight loss in obese individuals was associated with favourable hemodynamic effects,^9^ whereas a post-hoc analysis of the Prevención con Dieta Mediterránea (PREDIMED) trial showed a reduced risk of newly-diagnosed AF in those allocated to a nutritional intervention fostering the adherence to a Mediterranean Diet (MedDiet) enriched with extra-virgin olive oil in comparison with those who were allocated to the advice on a low-fat diet.^8^ Although the underlying pathophysiology mediating this effect is unknown, it seems plausible that lifestyle interventions can favourably reverse the remodelled atrial substrate or slow down its remodelling in people with increased weight and MetS, and therefore reduce their natural progression to AF.^10^ Investigations of such effects are important to validation of previous studies and personalization of therapies in at-risk individuals.

In this ancillary study of the PREDIMED-Plus trial, we aimed to evaluate the effect of an intensive lifestyle intervention (ILI) based on an energy-reduced MedDiet, increased physical activity, and cognitive-behavioural weight management on the underlying structural and functional cardiac substrate of AF in overweight or obese people with MetS.

## METHODS

### Study population

Participants were recruited for the PREvención con DIeta MEDiterranea-Plus (PREDIMED-Plus; ISRCTN89898870), which is a multicentre, randomised trial for the primary prevention of cardiovascular disease (CVD) in overweight/obese individuals with MetS. Between 2013 and 2016, 3,574 men (aged 55-75 years) and 3,300 women (aged 60-75 years) with a body mass index ≥27 and <40 kg/m^2^ meeting ≥3 criteria for the MetS were recruited from 23 centres in Spain. MetS was defined according to International Diabetes Federation, American Heart Association, and National Heart, Lung, and Blood Institute criteria.^11^ Participants were randomised 1:1 to an intensive lifestyle intervention (ILI) program based on an energy-reduced MedDiet, increased physical activity, and cognitive-behavioural weight management, or to a control intervention of low-intensity dietary advice on the MedDiet during at least 6 years. Primary endpoints of the trial are (1) a composite of non-fatal myocardial infarction, non-fatal stroke, or CVD death; and (2) weight loss and long-term weight-loss maintenance.

In this ancillary study of the PREDIMED-Plus trial, a sub-sample of 566 participants from 3 centres (University of Navarra-Preventiva, Hospital Universitario Araba, and Hospital Universitari Son Espases) were invited to evaluate the effect of the intervention on the underlying atrial substrate of AF. In this substudy, patients prospectively underwent TTE at baseline, 3-year and 5-year follow-up according to a uniform protocol, which has been detailed previously.^11^ After excluding participants with missing values in all TTE measures (n=20), and participants with baseline AF (n=12), 534 participants remained in the study population (**Figure 1**).

**Figure 1.**
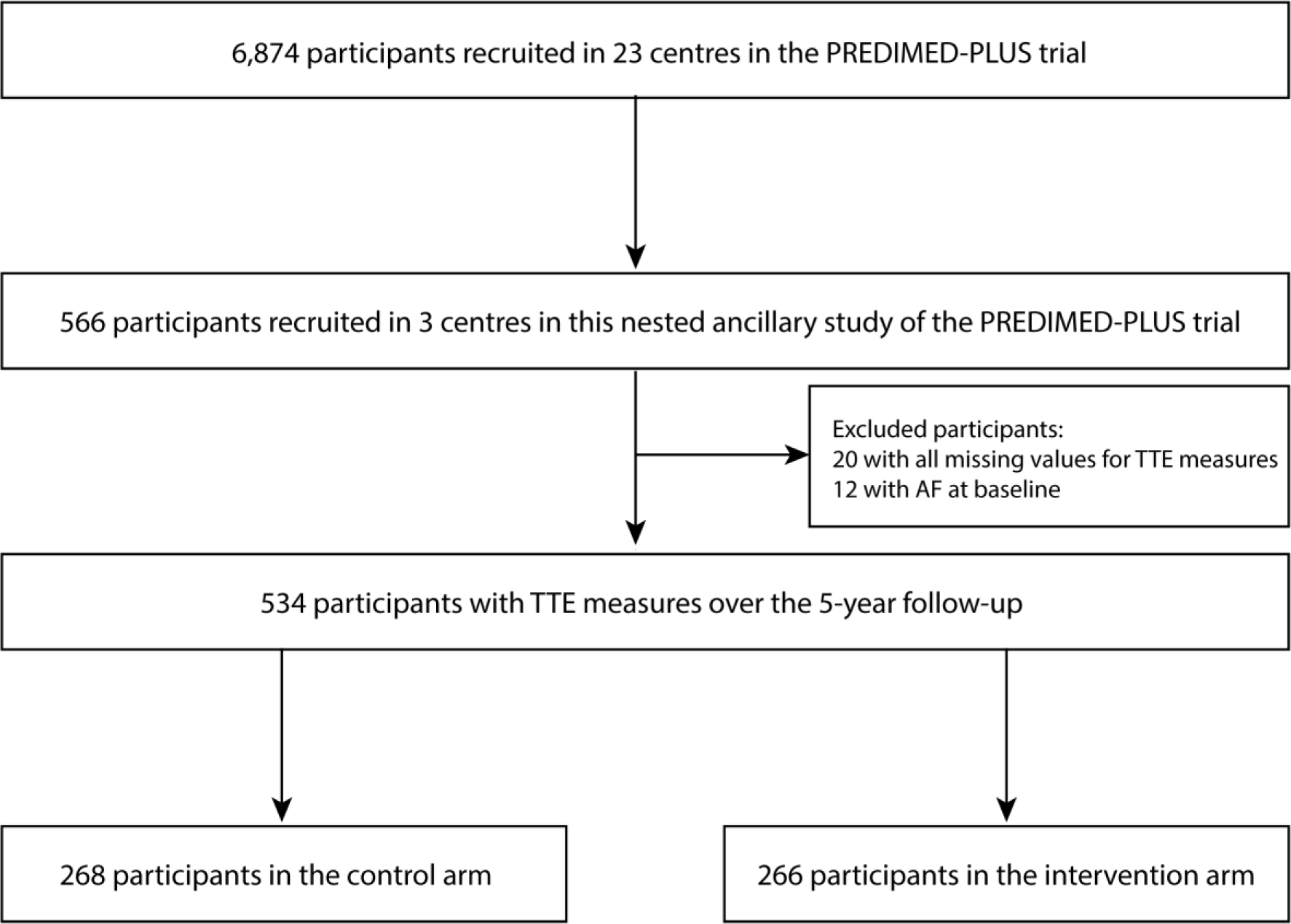
Study flow chart.

Images were then transferred to a core lab for their assessment.^12^ The rationale and design of the corelab Palma Echo Platform (PEP) has been published elsewhere.^12^.

### Study interventions

Participants randomised to the intervention group were instructed to follow an energy-reduced MedDiet, accompanied by physical activity promotion and behavioural support, with the purpose of accomplishing specific weight loss objectives.^13^ Trained dietitians conducted a group session, an individual motivational interview, and a phone call each month during the first year. Considering baseline customary levels of physical activity of each participant in the intervention group, a reduction of approximately 30% of estimated energy requirements (a reduction of roughly 600 kcal/d) was recommended. Participants in the control group attended 2 educational sessions per year on the traditional MedDiet without caloric restriction, all contents previously used in PREDIMED trial,^14^ and general lifestyle recommendations according to standard of care. Participants were also encouraged to progressively increase aerobic PA to ≥ 150 min/week of moderate-to-vigorous PA, with the final goal of walking ≥ 45 min/day or equivalent, and doing specific exercises to improve strength, balance, and flexibility. Compliance with PA recommendations was evaluated using validated questionnaires. Both groups of participants were supplied for free with extra-virgin olive oil (1 L/month) and all participants were encouraged to consume 500 g/month of mixed nuts.

### Study outcomes

TTE tests longitudinally and systematically assessed structural and functional features of the left atrium and other cardiac chambers. Data acquisition was performed according to the standards of the European Association of Echocardiography and the American Society of Echocardiography (ASE). ^15^ TTEs were performed at each site at baseline, 3-year and 5-year follow-up following a standardized protocol.^12^ Images were obtained at each centre by cardiologists with expertise in cardiac imaging, but they were assessed in a blind fashion by two independent expert cardiologists (core lab evaluators) not involved in obtaining the images. Each cardiologist was responsible for assessing a set of outcomes, so that each echocardiographic measure was assessed by one single cardiologist through the entire follow-up. The main study outcomes were LA emptying fraction, LA longitudinal strain –relative change in length of the tissue segment compared to its original length–, indexed and maximum LA volume, LA conduit strain –deformation in length of the LA during the passive filling phase of the cardiac cycle–, pump strain –deformation in length of the LA contraction or active emptying phase of the cardiac cycle–, LA stiffness index –as a measures of LA compliance–, and LA function index –overall performance of the LA–.^16–18^

### Other variables

In addition to the imaging data, some clinical information was collected, such as age, sex, cardiovascular risk factors and medical history at baseline. Anthropometric evaluations (weight, height, waist circumference) were measured according to the PREDIMED-Plus protocol. BMI was calculated as weight (kg) divided by the square of height (m^2^).

### Statistical analysis

These statistical analyses had two separate aims. The primary aim was to assess the effect of the ILI on the structural and functional cardiac substrate of AF, using TTEs tests performed at baseline, 3-year and 5-year follow-up. There were two primary outcomes: LA emptying fraction, and LA peak systolic longitudinal strain. Additionally, there were six secondary outcomes: indexed and maximum LA volume, LA conduit and pump strain, LA stiffness index, and LA function index.^17^ These outcomes collectively allowed us to measure both intermediate term functional effects that may be more dependent on real-time risk factor control, as well as longer-term structural effects that may improve more slowly over time in response to lifestyle change.^19^

The secondary aim was to estimate the association between changes in the 3 components of the ILI (adherence to the energy-reduced MedDiet, physical activity, and weight) and changes in echocardiographic measures at 3-year and 5-year follow-up. This allowed us to evaluate which specific component of the ILI may be the most impactful in reducing AF risk via LA substrate changes. We dichotomized the ILI changes to provide more clinical context/interpretability. According to the PREDIMED-Plus protocol, a clinically relevant weight loss was defined as a reduction in baseline body weight of over 8%, and a reduction in waist circumference of over 5%. For adherence to energy-reduced MedDiet, any improvement in the p17 questionnaire was defined as a change. For physical activity, achieving ≥150 min/week of moderate-to-vigorous exercise was defined as clinically meaningful. These changes were assessed over the 6 first months of follow-up.

Baseline characteristics were summarized as frequencies with percentages, or means with standard deviation (SD) and presented by study arm. We used mixed linear models to estimate differences in changes in LA measures from baseline to year 3 and to year 5 according to the intervention group. Time was modelled with 2 degrees of freedom (accounting for between-group differences in changes between year 3 and baseline and between year 5 and baseline). All models were adjusted for recruitment centre. We fitted a 2-level mixed linear model with random intercepts at cluster family and participant level. In an additional analysis, to address the relative changes in LA measurements over follow-up, we used linear mixed models with log-transformed LA measurements to assess the changes over time according to intervention group. Afterwards, estimates were back-transformed for an easier interpretation. As sensitivity analyses, we repeated the main analyses with multiple imputation methods using an iterative Markov chain Monte Carlo method (STATA “mi” command). We generated 20 imputations for each missing measurement. Imputed missing values were used for follow-up data but not for baseline data. Missing values for each outcome at baseline and follow-up are reported in **Supplementary Table 1**.^20^ To mitigate the potential impact of poor image quality, an additional analysis was conducted excluding participants with suboptimal echocardiographic windows. All analyses were conducted on an intention-to-treat basis.

To estimate the association of changes in the 3 components of the intervention (weight, adherence to the energy-reduced MedDiet, and physical activity) with changes in echocardiographic measures, a separate model was used to evaluate the impact of each individual component on LA longitudinal strain. In these models, the independent variables were the change (difference) between baseline and the 6-month post-randomisation visit in each one of the intervention components defined using the 17-item energy-reduced MedDiet questionnaire, weight in kilograms, waist circumference and physical activity based on the goals set to be achieved after 6 months in the trial protocol. For these analyses we used 2-level mixed linear models with random intercepts at cluster family and participant level adjusted for recruitment centre, intervention group, age, sex, prevalent diabetes, use of blood pressure lowering drugs (9 different drug families), and baseline systolic blood pressure. Ancillary, we also assessed weight, waist circumference, adherence to the energy-reduced MedDiet and time spent doing moderate-to-vigorous physical activity over follow-up according to intervention group, with 2-level mixed linear models with random intercepts at cluster family and participant level adjusted for recruitment centre.

All analyses were conducted using Stata version 16.1 (College Station, Texas, USA). A P-value of 0.05 was considered statistically significant. GraphPad Prism version 6.00 (GraphPad Software, La Jolla, California, USA) was used to produce the figures.

## RESULTS

### Baseline data

The number of patients included in the analysis is summarised in the flow chart shown in **Figure 1**. Baseline characteristics of the study participants according to their randomized group are presented in **Table 1**. Mean age at inclusion was 65 years (SD 5 years), and 40% of the participants were women. Given the inclusion criteria, prevalence of cardiovascular risk factors was high in both groups. At baseline, there were no substantial differences between groups regarding adherence to the MedDiet and total energy intake, though a higher physical activity amount was observed in the control as compared to the intervention arm. Regarding TTE baseline data, there were no major differences between groups (**Table 1**).

**Table 1.** Baseline clinical and echocardiographic data.

|  | Control<br>(n=268) | Intervention<br>(n=266) | Total in three<br>participating<br>sites<br>(n=534) | Total<br>PREDIMED<br>Plus<br>participants<br>(n=6874) |
| --- | --- | --- | --- | --- |
| **Baseline clinical data** |  |  |  |  |
| Women, % | 43 | 37 | 40 | 49 |
| Age, years | 66 (5) | 65 (5) | 65 (5) | 65 (5) |
| Body-mass index, kg/m<sup>2</sup> | 31.9 (3.2) | 32.5 (3.3) | 32.2 (3.3) | 32.6 (3.5) |
| Weight, Kg | 85.7 (12.4) | 88.5 (12.8) | 87.1 (12.7) | 86.6 (13.0) |
| Waist circumference | 105 (8) | 107 (8) | 106 (8) | 108 (10) |
| Smoking habit, % |  |  |  |  |
| Current smoker | 7 | 11 | 9 | 12 |
| Former smoker | 53 | 49 | 51 | 44 |
| Civil status, % |  |  |  |  |
| Married | 79 | 76 | 78 | 77 |
| Single | 6 | 5 | 6 | 5 |
| Widowed | 7 | 14 | 10 | 10 |
| Separated and divorced | 7 | 6 | 6 | 8 |
| Type 2 diabetes, % | 29 | 28 | 28 | 31 |
| Hypertension, % | 88 | 88 | 88 | 84 |
| Dyslipidemia, % | 74 | 75 | 74 | 70 |
| Physical activity, METs-min/d | 2833 (2269) | 2240 (2225) | 2538 (2265) | 2462 (2300) |
| Total energy intake, kcal/d | 2467 (712) | 2380 (614) | 2423 (666) | 2416 (633) |
| Adherence to the Mediterranean diet, 17-item screener | 7.6 (3.0) | 7.7 (2.9) | 7.6 (2.9) | 8.5 (2.7) |
| Alcohol intake, g/d | 17 (21) | 16 (21) | 16 (21) | 11 (15) |
| **Baseline echo data** |  |  |  |  |
| Baseline LA emptying fraction, % | 58.4 (9.4) | 59.4 (9.4) | 58.9 (9.4) | - |
| Baseline LA longitudinal strain, % | 27.5 (6.6) | 27.6 (6.5) | 27.6 (6.5) | - |
| Indexed LA volume, mL/m<sup>2</sup> | 22.7 (6.7) | 23.1 (7.4) | 22.9 (7.0) | - |
| LA volume, mL | 43.6 (14.0) | 45.0 (14.9) | 44.3 (14.5) | - |
| Conductile LA strain, % | -11.7 (4.7) | -12.2 (4.1) | -12.0 (4.4) | - |
| Contractile LA strain, % | -15.7 (4.6) | -15.6 (4.6) | 15.6 (4.6) | - |
| LA stiffness index | 0.3 (0.1) | 0.4 (0.2) | 0.4 (0.2) | - |
| LA function index | 67.1 (28.8) | 67.4 (29.6) | 67.3 (29.2) | - |
| LV ejection fraction, % | 65.7 (7.6) | 64.8 (5.8) | 65.2 (6.8) | - |
| LV longitudinal strain | -17.7 (2.6) | -17.7 (2.4) | -17.7 (2.5) | - |

LA, left atrium
LV, left ventricle

### Treatment effect on primary and secondary outcomes

Compared with the control group, the intervention group did not show a significant improvement in the primary outcomes (LA emptying fraction and LA longitudinal strain) over the 5-year period (**Figure 2**). Compared with baseline values, LA emptying fraction was lower at the end of follow-up for both groups (58.4% at baseline vs 56.4% at year 5 for the control arm, p=0.002; and 59.4% at baseline vs 57.7% at year 5 for the intervention arm, p=0.002), but not different between them (p_between groups_=0.80). Regarding LA longitudinal strain, there was a steady decline over time in both groups (27.5% at baseline vs 23.0% at year 5 for the control arm, p<0.001; and 27.6% at baseline vs 23.8% at year 5 for the intervention arm, p<0.001), but similarly there was no difference between groups (p_between groups_=0.24).

**Figure 2.**
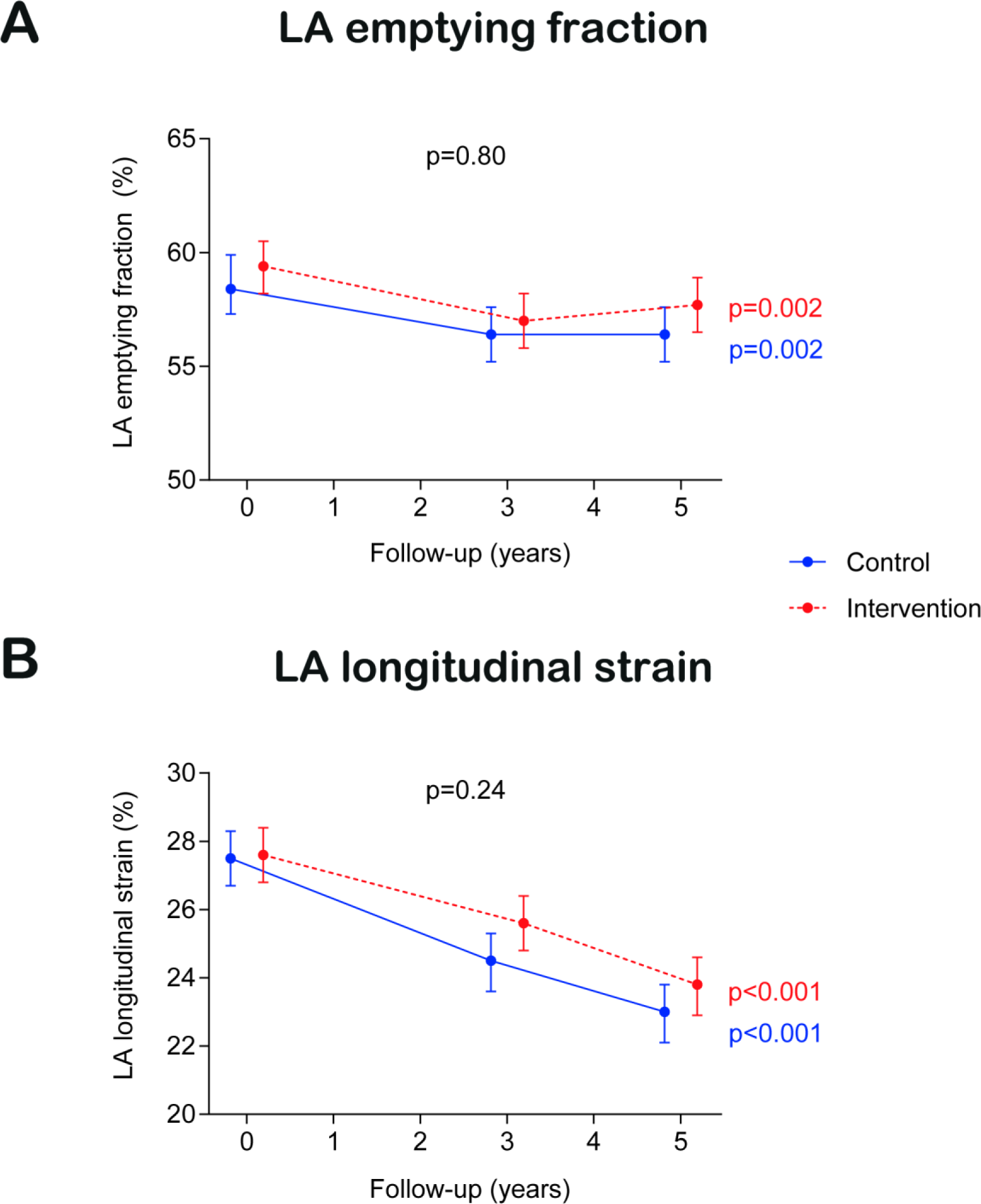
Treatment effect on co-primary outcomes. Results from linear mixed models. Time was modelled with 2 degrees of freedom. All models were adjusted for recruitment centre. We fitted a 2-level mixed linear model with random intercepts at cluster family and participant level.

The lack of treatment effect of the intervention was also observed across all the secondary outcomes (**Figure 3**): indexed LA volume (p_between groups_=0.72), maximum LA volume (p_between groups_=0.72), LA conduit strain (p_between groups_=0.12), LA pump strain (p_between groups_=0.31), LA stiffness index (p_between groups_=0.66), and LA function index (p_between groups_=0.99). For all endpoints, changes in endpoints followed a consistent trend in the control and the intervention group: indexed LA volume, maximum LA volume, and LA stiffness index increased over time, whereas LA conduit strain, LA pump strain, and LA function index decreased. Relative effects of the intervention on LA structural and functional echo parameters are shown as ratios of geometric means in **Table 2**, without evidence of a statistically significant effect of the intervention on any of the echocardiographic outcomes.

**Figure 3.**
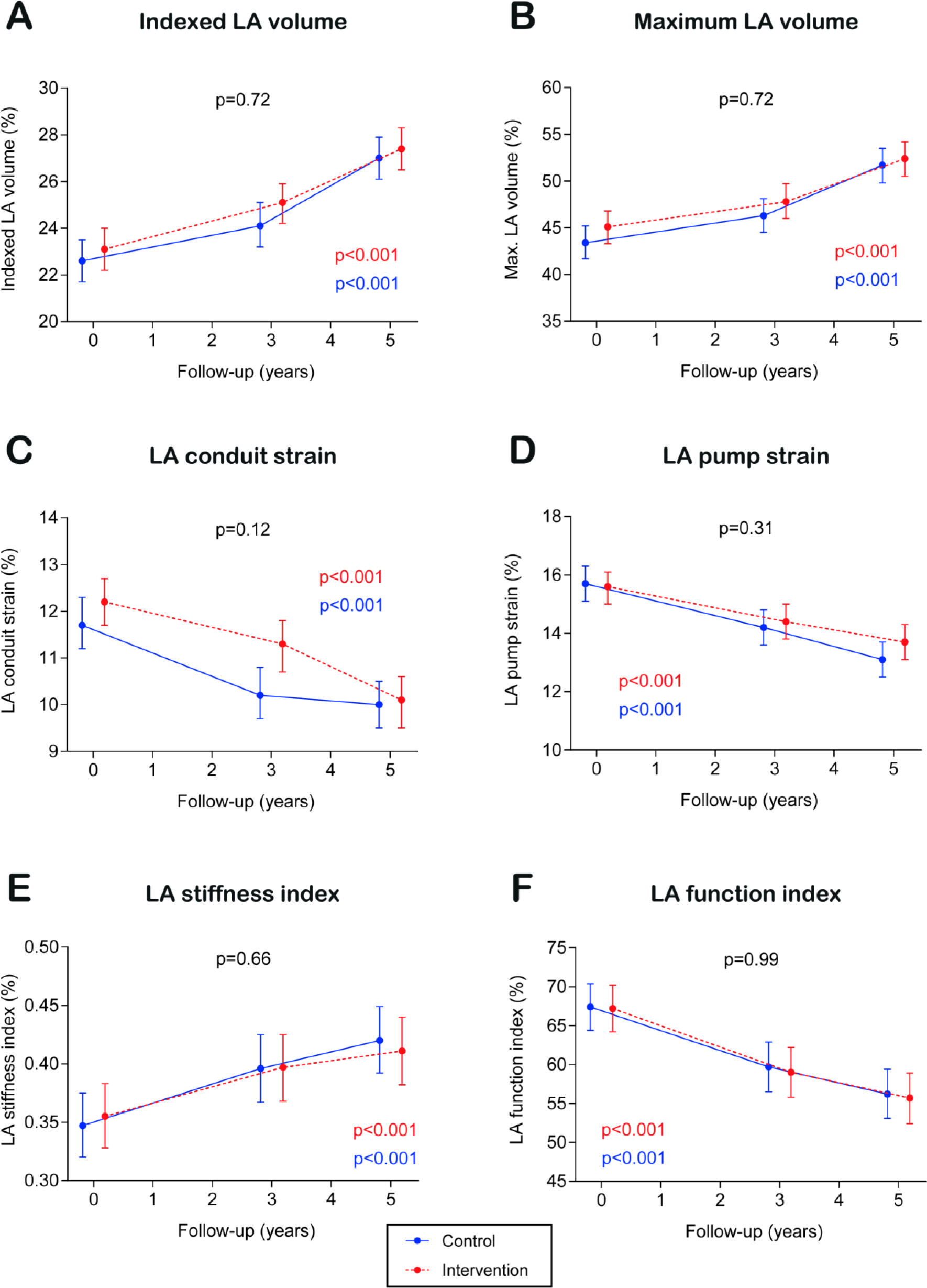
Treatment effect on secondary outcomes. Results from linear mixed models. Time was modelled with 2 degrees of freedom. All models were adjusted for recruitment centre. We fitted a 2-level mixed linear model with random intercepts at cluster family and participant level.

**Table 2.** Relative effect of an intensive lifestyle intervention on left atrium structural and functional parameters.

|  | Control group |  | ILI group |  | Difference ILI vs. control |  |
| --- | --- | --- | --- | --- | --- | --- |
|  | Y3 vs baseline | Y5 vs baseline | Y3 vs baseline | Y5 vs baseline | Y3 vs baseline | Y5 vs baseline |
| LA emptying fraction | 0.96 (0.94-0.99) | 0.95 (0.93-0.98) | 0.96 (0.93-0.98) | 0.97 (0.94-1.00) | 1.00 (0.96-1.03) | 1.01 (0.97-1.05) |
| LA longitudinal strain | 0.88 (0.85-0.92) | 0.82 (0.79-0.85) | 0.91 (0.88-0.95) | 0.85 (0.82-0.89) | 1.04 (0.98-1.09) | 1.04 (0.98-1.10) |
| Indexed LA volume | 1.07 (1.03-1.11) | 1.19 (1.15-1.23) | 1.09 (1.05-1.13) | 1.18 (1.14-1.23) | 1.02 (0.97-1.07) | 0.99 (0.94-1.05) |
| LA maximum volume | 1.07 (1.03-1.11) | 1.19 (1.14-1.23) | 1.07 (1.03-1.11) | 1.16 (1.12-1.20) | 1.00 (0.95-1.05) | 0.98 (0.93-1.03) |
| LA conduit strain | 0.88 (0.82-0.93) | 0.84 (0.79-0.89) | 0.91 (0.86-0.97) | 0.82 (0.77-0.87) | 1.04 (0.95-1.13) | 0.98 (0.90-1.06) |
| LA pump strain | 0.92 (0.87-0.97) | 0.84 (0.79-0.89) | 0.92 (0.87-0.97) | 0.88 (0.83-0.93) | 1.01 (0.93-1.09) | 1.05 (0.97-1.13) |
| LA stiffness index | 1.11 (1.05-1.17) | 1.17 (1.11-1.23) | 1.07 (1.02-1.13) | 1.10 (1.04-1.16) | 0.97 (0.90-1.04) | 0.94 (0.87-1.02) |
| LA function index | 0.89 (0.84-0.94) | 0.82 (0.78-0.87) | 0.88 (0.83-0.93) | 0.82 (0.78-0.87) | 0.99 (0.92-1.07) | 1.00 (0.92-1.08) |

Before performing mixed models, variables were log-transformed. Afterwards, they were back-transformed for an easier interpretation. In this table, values represent ratios of geometric means, alongside their 95% CI.
LA, left atrial

To evaluate the impact of missing values, the main analyses on primary and secondary outcomes were evaluated using a multiple imputation technique with 20 datasets (**Supplementary Table 2**). The findings for all outcomes were consistent to those yielded by the complete-case analysis.

To assess the influence of participants with suboptimal echocardiographic windows on the effects of ILI on primary outcomes, an additional analysis was conducted. The exclusion of these participants did not yield significant changes in the overall findings (**Supplementary Table 3**).

### Association between individual components of the intervention and LA echo parameters

The association of changes in components of the intervention (adherence to the energy-reduced MedDiet, physical activity, weight and waist circumference) with changes in echocardiographic measures was evaluated in all participants (**Figure 4**).

**Figure 4.**
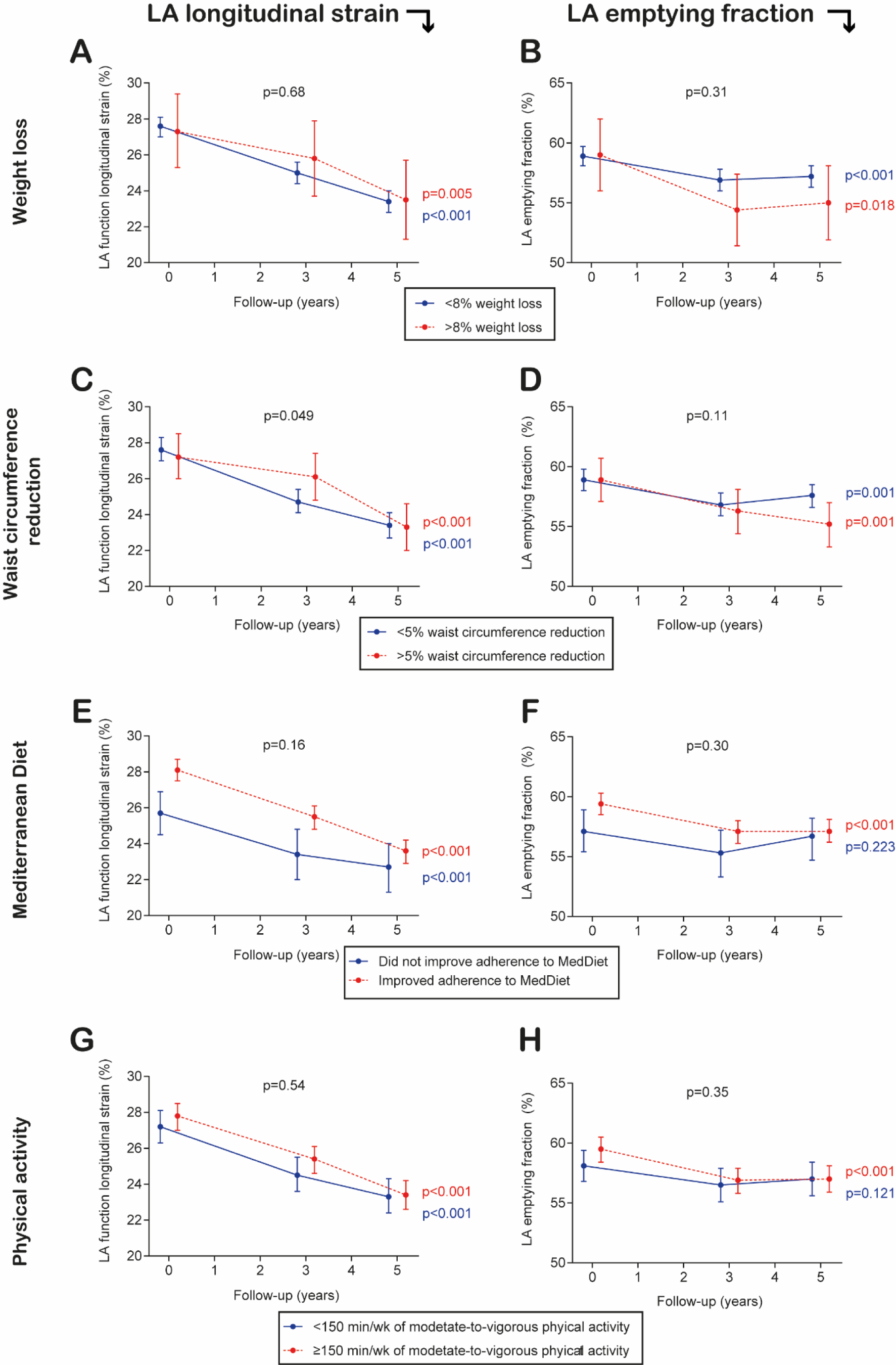
Association between individual components of the intervention and LA structural and functional parameters. Results from linear mixed models. Time was modelled with 2 degrees of freedom. All models were adjusted for recruitment centre. We fitted a 2-level mixed linear model with random intercepts at cluster family and participant level and adjusted for intervention group, age, sex, prevalent diabetes, use of blood pressure lowering drugs (9 different drug families), and baseline systolic blood pressure.

There were no significant differences in LA longitudinal strain over 5-year follow-up between those reducing and those not reducing >8% their weight loss (baseline vs. 5-year values for those not reducing weight loss were 27.6% vs 23.4%, whereas they were 27.3% vs 23.5% for those losing >8% weight; p_between groups_=0.68). There was a consistent decline in LA longitudinal strain over time in both groups (**Figure 4, panel A**). There were significant differences in weight over follow-up between the two interventions groups (Supplementary figure 1). Only participants in the ILI group significantly reduced their weight over follow-up. Weight loss took place during the first three years of the intervention and weight loss was sustained thereafter. In contrast, a borderline significant difference was observed when the change in waist circumference was evaluated (p_between groups_=0.049), though this difference was concentrated in the 3-year follow-up (**Figure 4, panel C**). There were significant differences in waist circumference over follow-up between the two interventions groups (Supplementary figure 2). Only participants in the ILI group significantly reduced their waist circumference over follow-up. As for weight, waist circumference reduction took place during the first three years of the intervention and weight loss was sustained thereafter.

There were no significant differences in LA longitudinal strain over 5-year follow-up between those who improved and those who did not improve the adherence to the energy reduced MedDiet (baseline vs. 5-year values for those not improving adherence were 25.7% vs 22.7%, whereas they were 28.1% vs 23.6% for those improving adherence to MedDiet; p_between groups_=0.16). There was a consistent decline in LA longitudinal strain in both groups between baseline and year 3, whilst in year 5 those not improving adherence did not have as lower values of LA longitudinal strain as those improving adherence (**Figure 4, panel E**). Both groups significantly increased their adherence to the energy-reduced MedDiet, although the changes were striker for the ILI group (Supplementary figure 3).

Regarding physical activity, there were no significant differences in LA longitudinal strain over 5-year follow-up between those who reached and those who did not reach the goal of ≥150 min/week of moderate-to-vigorous exercises (baseline vs. 5-year values for those not reaching goal were 27.2% vs 23.3%, whereas they were 27.7% vs 23.4% for those reaching that level of physical activity; p_between groups_=0.54; (**Figure 4, panel G**). There were significant differences in the time spent doing moderate-to-vigorous physical activity over follow-up between the two interventions groups (Supplementary figure 2). Only participants in the ILI group significantly increased their time spent doing moderate-to-vigorous physical activity over follow-up. This increase took place during the first three years of the intervention and weight loss was sustained thereafter.

The association between individual components of the intervention and LA emptying fraction was consistent with those observed for LA longitudinal strain (**Figure 4, panel B, D, F, H**).

## DISCUSSION

In this nested randomised clinical trial, conducted in overweight or obese people with MetS, an ILI program based on an energy-reduced MedDiet, increased physical activity, and cognitive-behavioural weight management did not show a significant impact on the structural and functional cardiac substrate of AF compared a control intervention of low-intensity dietary advice on the MedDiet. There were significant changes in all echocardiographic endpoints (LA emptying fraction, LA peak systolic longitudinal strain, indexed and maximum LA volume, LA conduit and pump strain, LA stiffness index, and LA function index) in both arms, though these changes were similar by arm. There were no significant differences in LA longitudinal strain and LA emptying EF change based on the intervention components of weight loss, improvement in adherence to MedDiet, and physical activity. Compared to those with less waist circumference reduction, LA longitudinal strain decreased slower in those with the highest waist circumference reduction at year 3, although this was not observed at year 5.

People with MetS and increased body weight are at high risk for AF development and progression, since these conditions have an effect on cardiovascular hemodynamics (e.g., hypertension), as well as a consequent long-term impact on cardiac function and structure.^5,6^ Some changes in LA, such as a greater increase in LA volumes and decrease in LA function, have been considered a surrogate outcome for developing AF.^10^ Our findings do not support that an ILI plays a role in preventing LA changes, though these new results need to be put in context. Previous randomised and observational studies summarised in a meta-analysis showed that weight loss in obese individuals was associated with favorable hemodynamic effects,^9^ which can be considered the preliminary step before changes in LA take place. A post-hoc analysis of the PREDIMED trial showed that an intervention promoting adherence to the MedDiet enriched with extra-virgin olive oil can reduce by 40% the risk of newly-diagnosed AF compared with a control intervention promoting a low-fat diet. Our data do not contradict the findings yielded by these studies, since we do not report neither the hemodynamic changes nor the incidence of AF over time. However, our results do not support that our intervention, an ILI program based on an energy-reduced MedDiet, increased physical activity, and cognitive-behavioural weight management, have an impact on the changes that happen on LA characteristics over time in overweight or obese people with MetS, except for waist circumference at 3 years on LA strain. Visceral adiposity is likely the be one of the most important risk factors for AF, and longitudinal strain may be superior to the other measures in terms of its ability to reflect improved health status and predict AF risk.^21^ Other mechanisms, such as autonomic or electrocardiographic, warrant further examination.^22,23^

Several reasons might explain the lack of treatment effect provided by an ILI. Given that the control intervention was a dietary advice on the MedDiet, it is reasonable to think that the “active comparator” might have had already an impact on LA changes in the control group.^8^ It might also happen that the ILI is not having an impact on LA outcomes, unlike other lifestyle interventions might have. This is supported by the data provided for the attainment of each individual component of the intervention. Given that those patients with higher degree of compliance and achievements of clinically relevant goals (weight loss, MedDiet adherence, physical activity) did not have different outcomes than those with a lower degree of compliance, it seems that none of the individual interventions is driving any treatment effect. We tested an ILI under the hypothesis that smaller changes in LA will result in a reduced number of AF episodes in the long-term. This paradigm assumes that the risk for AF is greatly dependent on the degree of LA impairment. Eventually, LA changes could be good clinical markers, but not the best surrogate endpoints. Nevertheless, in the MESA study, a cohort of people free of CVD at baseline, a greater increase in LA volumes and decrease in LA function was associated with incident AF, whereas a change in LA emptying fraction improved model discrimination and reclassification of AF risk.^10^ Finally, intervention effects on lifestyle might have attenuated over time, suggesting that sustained effects need a different strategy to change behaviours in the long term. There are several examples of a transient favourable change after a lifestyle intervention that tend to dilute over time, such as those observed in the recently published TANSNIP-PESA (Trans-Atlantic Network to study Stepwise Non-invasive Imaging as a tool for CVD Prognosis and prevention) study.^24^ Ancillary analyses conducted for this subsample showed that the lifestyle intervention led to changes in its individual components during the first three years of follow-up and were sustained thereafter. In addition, the COVID-19 pandemic might have had an impact of the trial conduct, though no major effects have been observed in previous reports.^25^ Our intervention has effect on some biomarkers that are also surrogates for AF,^26^ but might have little influence in echo parameters should they be already altered before enrolment in the trial (e.g., long-standing obesity).^27^ There were no significant differences in LA longitudinal strain and LA emptying fraction according to changes in each individual component of the intervention, which was consistent with the lack of treatment effect on the primary outcomes.

Both AF and metabolic syndrome (MetS) are becoming a global public health issue due to the aging of the population and the increasing levels of obesity and physical inactivity among adults in many countries.^28,29^ Although we failed to show efficacy on echocardiographic outcomes in the comparison between ILI and low-intensity dietary advice on the MedDiet, it seems plausible that other lifestyle interventions can favourably reverse the remodelled cardiac substrate of people with increased weight and MetS, and therefore reduce their natural progression to AF.^10^ In the REVERSE-AF (PREVEntion and regReSsive Effect of weight-loss and risk factor modification on Atrial Fibrillation) study,^3^ weight loss and risk factor management reversed the type and natural progression of AF in patients with already some form of AF. Similarly, aggressive risk factor reduction improved the long-term success of AF ablation, underscoring the importance of therapy directed at the primary promoters of the AF substrate to facilitate rhythm control strategies.^30^

The use of echocardiography in this setting deserves special attention. The atrium has been historically evaluated in its morphological dimension (e.g., atrial volume). In our study, we evaluated novel parameters, such as LA strain and emptying fraction.^31,32^ Unlike traditional parameters based on blood flow during atrial contraction such as the peak A wave velocity or the A wave velocity time integral, LA strain represents a reliable direct measure of atrial function.^31,32^

### Strengths and limitations of this study

Strengths of this trial include an adequate sample size to fairly answer the main hypothesis, the randomised design, reducing the threat of confounding, the prolonged intervention and follow-up periods for the study of lifestyle-change maintenance, the excellent retention during follow-up, and the objective measurement of the echo outcomes. Nesting this study within the PREDIMED-PLUS trial provides a perfect opportunity for further comprehensive assessments of the association between lifestyle changes and myocardial derangements. Nevertheless, several limitations should be considered. This is a nationwide trial of highly selected patients (increased weight with MetS enriched with at least 3 factors), and therefore the generalisation of the findings should be taken cautiously.^33^ Due to the type of intervention, neither participants nor investigators were blinded to the allocation arm, though we were masked for the statistical analysis and all study outcomes relied on objective parameters not subject to self-report bias. The level of statistical significance has not been adjusted for the number of comparisons^34^. It should be acknowledged that an optimal TTE acquisition is particularly challenging in a population with a high body mass index and MetS. It is also challenging to obtain standard values in a hypothetical reference population, given the difference between commercial software’s measuring LA strain. Other techniques, such as cardiac magnetic resonance, might be useful to complement echo measures in future studies. Finally, the long-term overall results for risk factor outcomes were outside of the scope of this analysis, nevertheless significant changes in major cardiovascular risk factors after one year of follow-up have been observed.^13^

## CONCLUSIONS

Using randomised data from overweight or obese people with MetS, we observed that an ILI program based on an energy-reduced MedDiet, increased physical activity, and cognitive-behavioural weight management did not have an effect on none of the primary (LA emptying fraction, and LA peak systolic longitudinal strain) or secondary (indexed and maximum LA volume, LA conduit and pump strain, LA stiffness index, and LA function index) echocardiographic endpoints. Our findings suggest that there are age related changes over time in these parameters that indicate increased AF risk, but that an ILI has no impact on them. Hence, an ILI had no impact on the underlying structural and functional cardiac substrate of AF in overweight or obese people with MetS. Similarly, those achieving higher goals in terms of weight loss, adherence to MedDiet, or physical activity, showed no difference in LA longitudinal strain compared to those below clinically relevant changes.

## Data Availability

There are restrictions on the availability of data for the PREDIMED-Plus study, due to the signed consent agreements around data sharing, which only allow access to external researchers for research following the project purposes. Requestors wishing to access the PREDIMED-Plus trial data used in this study can request it to the PREDIMED-Plus trial Steering Committee.

## ACKNOWLEDGMENTS

The authors especially thank the PREDIMED-Plus participants for the enthusiastic collaboration, the PREDIMED-Plus personnel for outstanding support, and the personnel of all associated primary care centers and cardiology departments for the exceptional effort.

## SUPPLEMENTARY TABLES

**Supplementary Table 1.** Participants with missing values in the baseline and follow-up ascertainment of the primary and secondary outcomes.

| Outcome | Baseline | Follow-up |
| --- | --- | --- |
| LA emptying fraction, n | 2 | 32 |
| LA longitudinal strain, n | 7 | 33 |
| Indexed maximum LA volume | 2 | 35 |
| Maximum LA volume | 2 | 32 |
| LA conduit and pump strain | 24 | 33 |
| LA stiffness index | 8 | 35 |
| LA function index | 12 | 35 |

LA, left atrial.

**Supplementary Table 2.** Sensitivity analyses of the effect of an intensive lifestyle intervention on left atrium structural and functional parameters using multiple imputations for missing data.

|  | Control | Intervention | p-value<br>(within groups) | p-value (between<br>groups) |
| --- | --- | --- | --- | --- |
| LA EF |  |  |  |  |
| Baseline, mean (95% CI) | 58.5 (57.4-59.7) | 59.3 (58.2-60.5) | P<sub>int</sub>=0.002<br>P<sub>con</sub>=0.003 | 0.80 |
| Year 3, mean (95% CI) | 56.5 (55.2-57.7) | 57.0 (55.8-58.2) |  |  |
| Year 5, mean (95% CI) | 56.4 (55.2-57.7) | 57.6 (56.4-58.9) |  |  |
| LA longitudinal strain |  |  |  |  |
| Baseline, mean (95% CI) | 27.6 (26.8-28.4) | 27.6 (26.8-28.4) | P<sub>int</sub><0.001<br>P<sub>con</sub><0.001 | 0.24 |
| Year 3, mean (95% CI) | 24.5 (23.7-25.4) | 25.6 (24.7-26.4) |  |  |
| Year 5, mean (95% CI) | 23.0 (22.1-23.9) | 23.8 (22.9-24.6) |  |  |
| Indexed LA volume |  |  |  |  |
| Baseline, mean (95% CI) | 22.6 (21.7-23.4) | 23.1 (22.3-24.0) | P<sub>int</sub><0.001<br>P<sub>con</sub><0.001 | 0.72 |
| Year 3, mean (95% CI) | 24.1 (23.2-25.0) | 25.1 (24.2-26.0) |  |  |
| Year 5, mean (95% CI) | 26.9 (26.0-27.9) | 27.4 (26.5-28.4) |  |  |
| Maximum LA volume |  |  |  |  |
| Baseline, mean (95% CI) | 43.4 (41.6-45.2) | 45.2 (43.4-47.0) | P<sub>int</sub><0.001<br>P<sub>con</sub><0.001 | 0.75 |
| Year 3, mean (95% CI) | 46.2 (44.4-48.1) | 47.9 (46.1-49.8) |  |  |
| Year 5, mean (95% CI) | 51.6 (49.7-53.4) | 52.5 (50.6-54.3) |  |  |
| LA conduit strain |  |  |  |  |
| Baseline, mean (95% CI) | 11.8 (11.3-12.3) | 12.1 (11.6-12.7) | P<sub>int</sub><0.001<br>P<sub>con</sub><0.001 | 0.12 |
| Year 3, mean (95% CI) | 10.3 (9.7-10.8) | 11.3 (10.7-11.8) |  |  |
| Year 5, mean (95% CI) | 10.0 (9.5-10.6) | 10.1 (9.5-10.6) |  |  |
| LA pump strain |  |  |  |  |
| Baseline, mean (95% CI) | 15.7 (15.1-16.3) | 15.6 (15.0-16.2) | P<sub>int</sub><0.001<br>P<sub>con</sub><0.001 | 0.31 |
| Year 3, mean (95% CI) | 14.2 (13.6-14.8) | 14.4 (13.8-15.0) |  |  |
| Year 5, mean (95% CI) | 13.1 (12.5-13.7) | 13.7 (13.1-14.3) |  |  |
| LA stiffness index |  |  |  |  |
| Baseline, mean (95% CI) | 0.35 (0.32-0.37) | 0.36 (0.33-0.38) | P<sub>int</sub><0.001<br>P<sub>con</sub>=0.001 | 0.65 |
| Year 3, mean (95% CI) | 0.40 (0.37-0.42) | 0.40 (0.37-0.43) |  |  |
| Year 5, mean (95% CI) | 0.42 (0.39-0.45) | 0.41 (0.38-0.44) |  |  |
| LA function index |  |  |  |  |
| Baseline, mean (95% CI) | 67.5 (64.4-70.5) | 67.2 (64.2-70.2) | P<sub>int</sub><0.001<br>P<sub>con</sub><0.001 | 0.96 |
| Year 3, mean (95% CI) | 59.9 (56.7-63.1) | 58.9 (55.7-62.0) |  |  |
| Year 5, mean (95% CI) | 56.3 (53.1-59.5) | 55.6 (52.4-58.9) |  |  |

LA, left atrial

**Supplementary Table 3.** Relative effect of an intensive lifestyle intervention on left atrium structural and functional parameters among visits with good echocardiographic quality.

|  | Control group |  | ILI group |  | Difference ILI vs. control |  |
| --- | --- | --- | --- | --- | --- | --- |
|  | Y3 vs baseline | Y5 vs baseline | Y3 vs baseline | Y5 vs baseline | Y3 vs baseline | Y5 vs baseline |
| LA emptying fraction | 0.96 (0.93-0.99) | 0.93 (0.91-0.96) | 0.96 (0.93-0.98) | 0.95 (0.93-0.98) | 1.00 (0.96-1.04) | 1.02 (0.98-1.06) |
| LA longitudinal strain | 0.92 (0.88-0.96) | 0.82 (0.79-0.86) | 0.94 (0.90-0.97) | 0.87 (0.84-0.91) | 1.02 (0.96-1.08) | 1.06 (1.00-1.12) |
| Indexed LA volume | 1.06 (1.02-1.11) | 1.22 (1.17-1.27) | 1.07 (1.03-1.11) | 1.18 (1.13-1.23) | 1.01 (0.95-1.07) | 0.97 (0.91-1.03) |
| LA maximum volume | 1.06 (1.02-1.11) | 1.21 (1.16-1.26) | 1.04 (1.00-1.09) | 1.15 (1.11-1.20) | 0.98 (0.93-1.04) | 0.95 (0.90-1.01) |
| LA conduit strain | 0.90 (0.84-0.96) | 0.84 (0.79-0.90) | 0.93 (0.87-0.99) | 0.85 (0.80-0.91) | 1.03 (0.95-1.13) | 1.02 (0.93-1.12) |
| LA pump strain | 0.96 (0.91-1.02) | 0.86 (0.81-0.92) | 0.95 (0.90-1.01) | 0.91 (0.86-0.96) | 0.99 (0.91-1.08) | 1.05 (0.97-1.14) |
| LA stiffness index | 1.07 (1.01-1.14) | 1.16 (1.09-1.23) | 1.05 (0.99-1.11) | 1.08 (1.01-1.14) | 0.98 (0.90-1.07) | 0.93 (0.85-1.01) |
| LA function index | 0.91 (0.85-0.97) | 0.79 (0.74-0.85) | 0.91 (0.85-0.97) | 0.81 (0.76-0.86) | 1.00 (0.91-1.10) | 1.02 (0.93-1.12) |

Before performing mixed models, variables were log-transformed. Afterwards, they were back-transformed for an easier interpretation. In this table, values represent ratios of geometric means, alongside their 95% CI.
LA, left atrial

**Supplementary figure 1.**
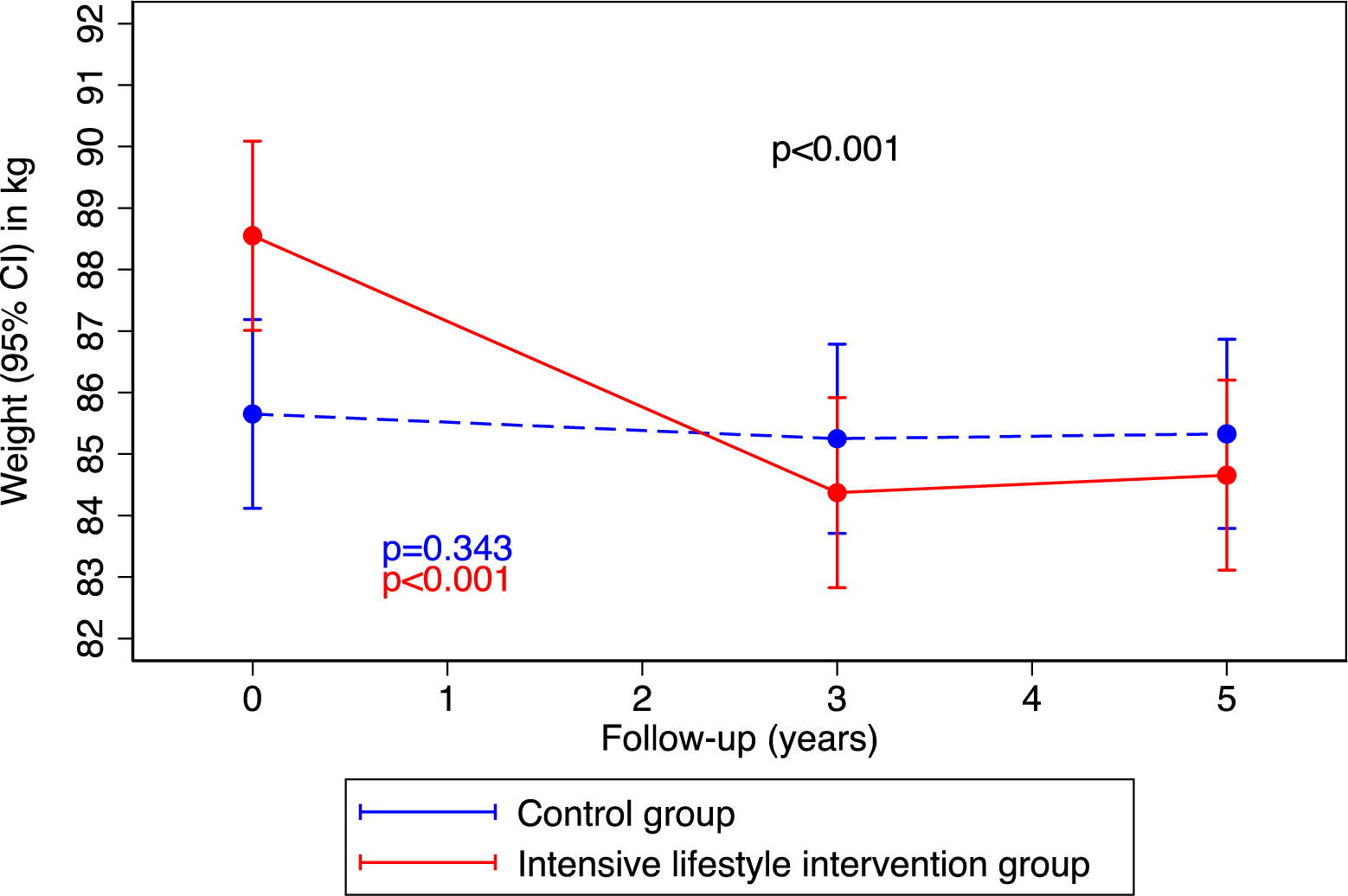
Weight over follow-up according to the intervention group. Results from linear mixed models. Time was modelled with 2 degrees of freedom. All models were adjusted for recruitment centre. We fitted a 2-level mixed linear model with random intercepts at cluster family and participant level.

**Supplementary figure 2.**
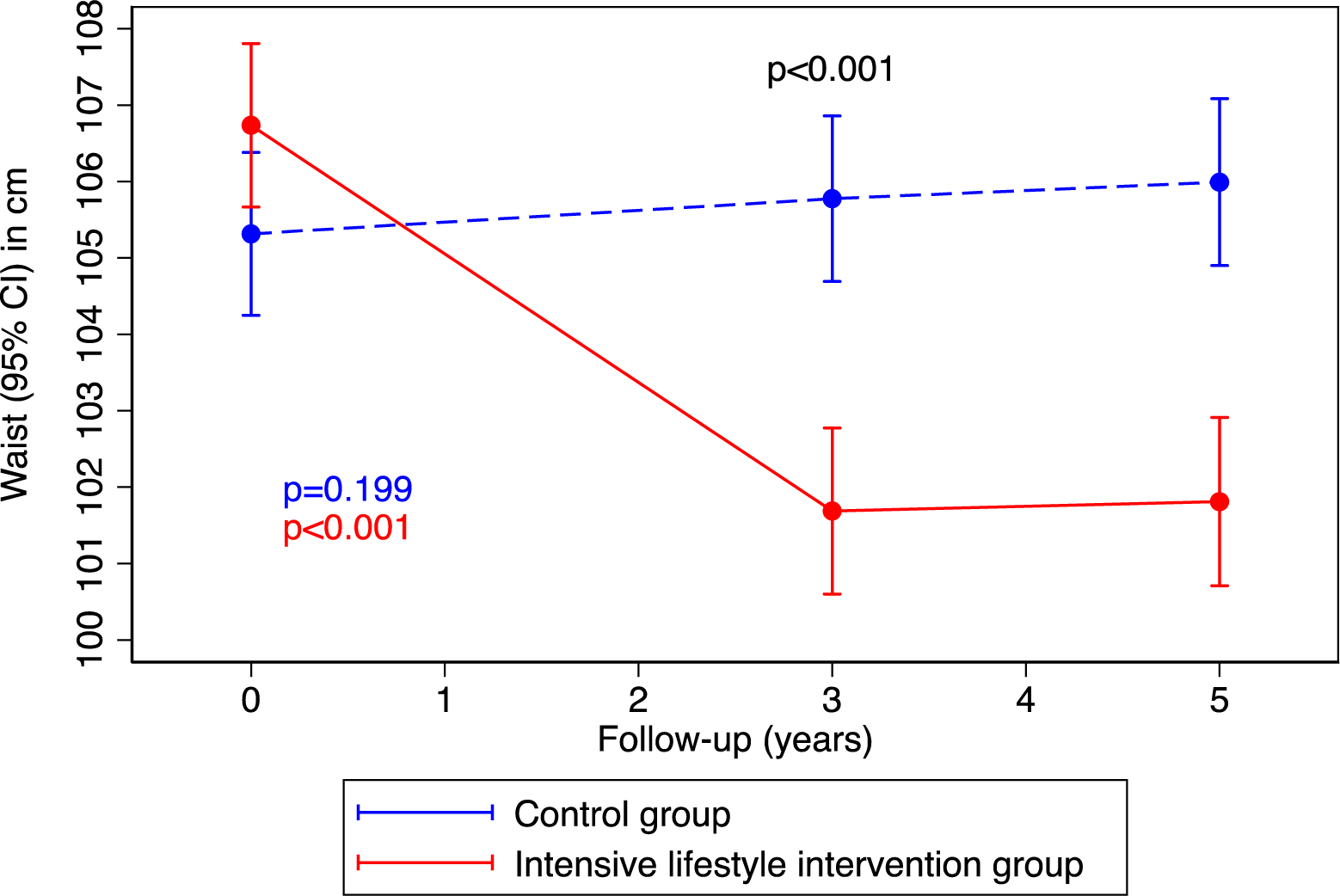
Waist circumference over follow-up according to the intervention group. Results from linear mixed models. Time was modelled with 2 degrees of freedom. All models were adjusted for recruitment centre. We fitted a 2-level mixed linear model with random intercepts at cluster family and participant level.

**Supplementary figure 3.**
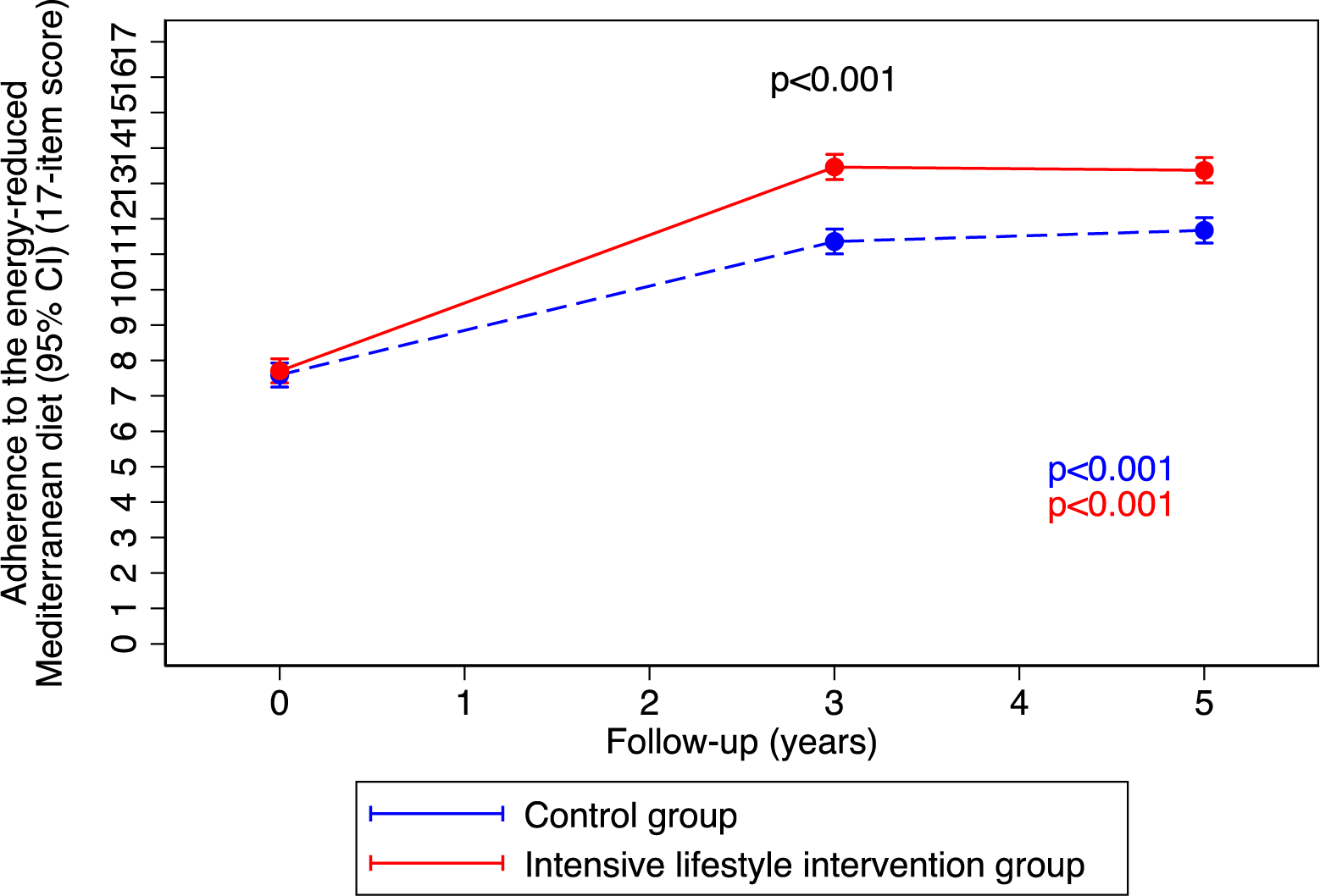
Adherence to the energy-reduced Mediterranean diet over follow-up according to the intervention group. Results from linear mixed models. Time was modelled with 2 degrees of freedom. All models were adjusted for recruitment centre. We fitted a 2-level mixed linear model with random intercepts at cluster family and participant level.

**Supplementary figure 4.**
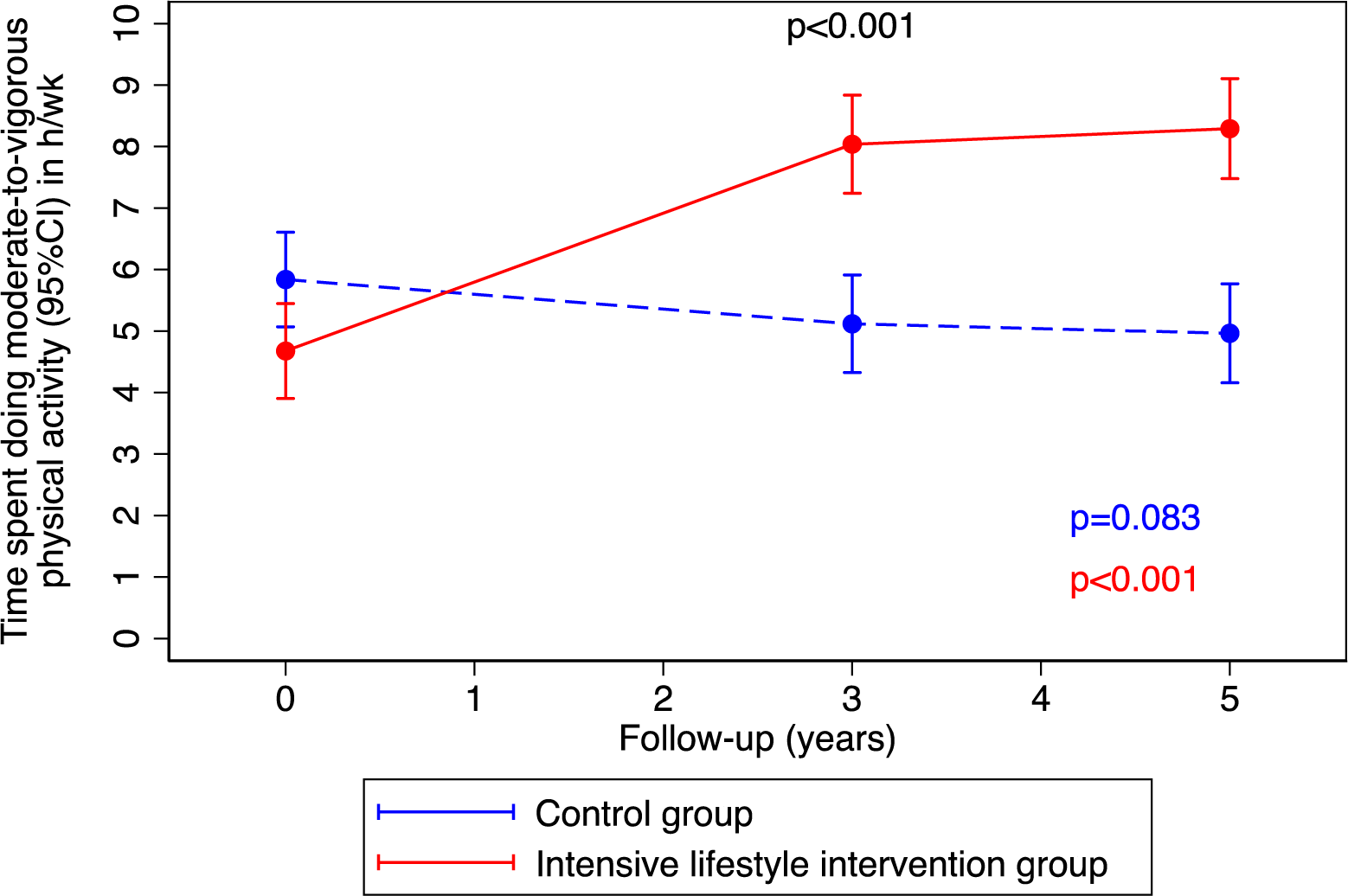
Time spent doing in moderate-to-vigorous physical activity over follow-up according to the intervention group. Results from linear mixed models. Time was modelled with 2 degrees of freedom. All models were adjusted for recruitment centre. We fitted a 2-level mixed linear model with random intercepts at cluster family and participant level.

